# Assessment of Elapsed Time Between Dental Radiographs Using Siamese Network

**DOI:** 10.1101/2025.03.04.25323305

**Authors:** Marija Milutinovic, René Daher, Julian Leprince, Douglas Teodoro

**Affiliations:** University of Geneva, Faculty of Medicine, Medical Informatics and Radiology; University of Geneva, University Clinics of Dental Medicine, Division of Cariology and Endodontology

**Keywords:** deep learning, dental radiographs, time span prediction

## Abstract

Recently, machine learning methods have emerged to predict dental disease progression, often relying on costly annotated datasets and frequently exhibiting low generalization performance. This study evaluates the application of Siamese networks for detecting subtle changes in longitudinal dental x-rays and predicting the time span category between dental treatments using periapical radiographs and patient demographic data. We assume that the ability of these models to detect the time intervals between dental treatments would ensure their capability to identify more complex patterns related to disease progression. The baseline models based on CNNs and MLP achieved moderate performance, while the Siamese network models demonstrated significant improvements, with the highest-performing model achieving an accuracy of 86.32% ± 1.60%. Moreover, the introduction of demographic features such as age and gender into the model led to a significant reduction in performance variance. These results underscore the effectiveness of Siamese networks in capturing subtle temporal changes in dental radiographs in longitudinal settings, offering the potential to integrate these models into clinical workflows. Future research will explore self-supervised learning models for dental disease progression, especially in clinical settings with limited labeled data.

## 1. Introduction

Dental radiographs are necessary tools for diagnosing and treating oral diseases, allowing dentists to visualize structures not visible through direct examination [1,2]. The conventional analysis of these radiographs involves manual interpretation, which can be time-consuming and susceptible to human error. This is particularly the case for assessing risks of dental disease progression in early stages, such as caries, bone loss, periodontitis, and eventually tooth loss, when changes between longitudinal radiographs might be too subtle to the human eye [3]. As a complementary approach, recent advancements in deep learning techniques have shown promise in enhancing the accuracy and efficiency of image analysis in dentistry [2,4].

Deep learning methods for risk assessment of periodontally compromised teeth use longitudinal information to provide early decision support to dentists [5]. By training on annotated data about disease progression, these algorithms can extract patterns from early-stage data, enabling more accurate risk estimation [6]. Previously, machine learning models such as LASSO and K-means were employed to predict mandibular growth and caries risk over time [7,8] using features extracted from dental images. While these methods have reached promising performance, the need for large annotated dental X-ray datasets for model training hinders their assessment in terms of generalizability and thus application in real-world clinical decision support scenarios [9].

Instead of estimating risks of disease progression, we simplify the task and focus on the assessment of time span between longitudinal dental radiographs as a paramount requirement for biological disease progression. Using a dataset of 470 periapical radiographs and demographic information, longitudinal instances belonging to the same patient were paired and labeled within 3 time bins: less than or equal to 1 year, more than 1 year and less than or equal to 5 years, or more than 5 years. We then explored the application of the Siamese network, a neural network designed to learn similarities between input pairs [6,9], to predict time intervals between longitudinal pairs of patient data. We report on the model’s classification performance and on the visual features used to predict the time difference between radiographs.

## 2. Method

### 2.1. Dataset description

The raw dataset used in this study consists of periapical radiographs and demographic information from 146 patients. Each patient’s record includes an anonymized ID, date of birth, gender, treatment start and end dates, treated tooth, and a series of radiographs obtained across multiple dental treatments, along with an indication of whether a digital radiograph is available. The benchmark dataset was constructed by pairing radiographs from two consecutive patient visits and the corresponding elapsed time between treatments was calculated, resulting in 470 samples. Patient age was recorded at the start of treatment and categorized into one of ten age groups, ranging from 0 to 90 years, with each group spanning 10 years. Both age and gender were treated as categorical features and used as inputs to the model. The demographic data is presented in Table 1. The use of the data for retrospective analysis was approved by the ethical commitee of the Cliniques universitaires Saint-Luc, Brussels, Belgium (#2024/12NOV/492).

**Table 1.** Statistics for the dataset used in the experiment.

| Elapsed time<br>(group 0) | time ≤ 1 year<br>(group 1) | 1 year < time ≤ 5 years<br>(group 2) | time > 5 years |
| --- | --- | --- | --- |
| Number of samples | n = 88 | n = 284 | n = 98 |
| Gender [male/female] | 54/34 | 183/101 | 60/38 |
| Age (years) | 47.8 ± 14.5 | 50.8 ± 13.8 | 51.8 ± 11.9 |
| Beginning of treatment | 1996-09-16 | 1994-10-04 | 1992-09-16 |
| End of treatment | 2015-12-21 | 2015-08-18 | 2010-09-21 |
| Average elapsed time [Years] | 3.20 ± 2.48 | | |

### 2.2. Architecture

To predict temporal intervals between dental treatments using periapical radiographs, we employed a Siamese network architectures. Siamese networks are characterized by their ability to learn the similarity between pairs of inputs [10,11] and are particularly effective for tasks that involve comparisons [12], such as determining the time intervals between radiographic images. As shown in Figure 1, the Siamese network architecture utilized in this study comprises two identical sub-networks. Each sub-network processes one of the input images through a series of convolutional layers that extract hierarchical features, followed by fully connected dense layers that produce a compact feature representation. The outputs of these sub-networks are then compared using a distance metric, typically the Euclidean distance, to quantify the similarity between the two images. This similarity score is subsequently used to estimate the time elapsed between the radiographs.

**Figure 1.**
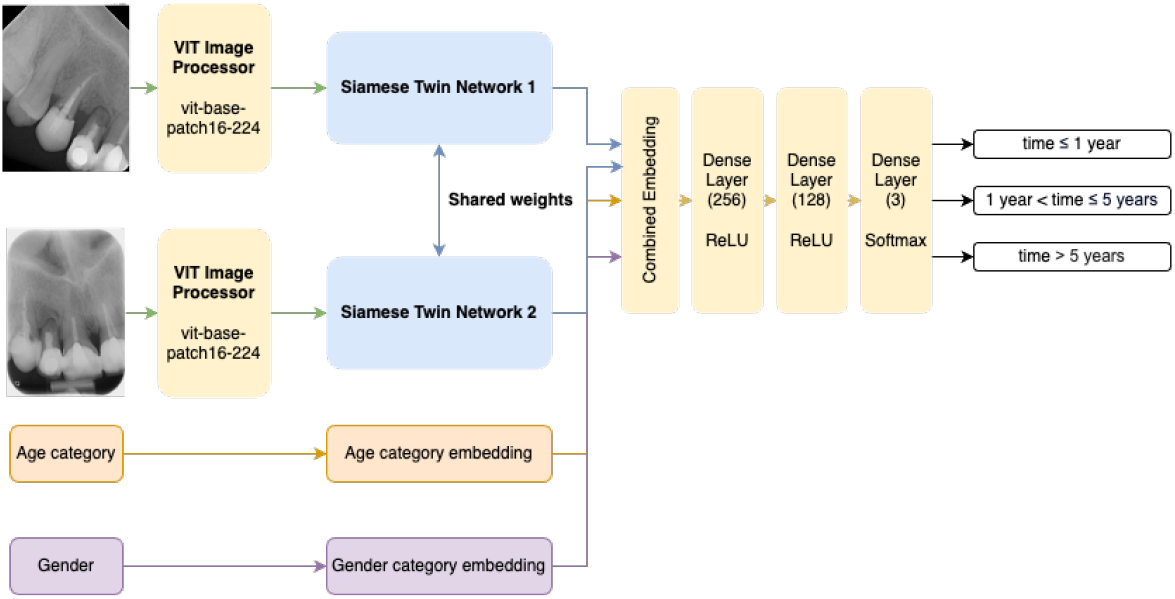
The Siamese network architecture with multimodal features.

The raw radiographs were preprocessed using google/vit-base-patch16-224 ViTImageProcessor. The training process employed a contrastive loss function, which minimizes the distance between feature representations of radiograph pairs taken close in time while maximizing the distance for pairs taken further apart. The Adam optimizer was used with an initial learning rate of 0.001, and the network was trained for 100 epochs with a batch size of 16, and the margin for constrastive loss was set to 1. During training, a validation set was monitored to prevent overfitting, and hyperparameter tuning for batch size, learning rate and optimizer was performed using Optuna.

### 2.3. Model training and evaluation

Model was trained using 5-fold cross-validation on 80% of benchamark dataset and evaluated on hold-out test set for each fold. Three variants of the Siamese network were tested: a basic model with standard convolutional and dense layers and categorical cross-entropy, an enhanced model with constrastive loss, and final model that leverages multimodal data. The Siamese models’ performance was compared to baseline models based on traditional convolutional neural networks and multi-layer perceptron. We interpret the best model’s prediction using Grad-CAM (Gradient-weighted Class Activation Mapping), which highlights parts of the input image which contribute the most to a specific prediction [13].

## 3. Results and Discussion

The performance of the models to predict time spans between periapical radiographs is presented in Table 2. The Siamese model, using multimodal data, transfer learning via visual transformers, and contrastive loss, achieved the highest performance, with and accuracy of 86.32% and F1 score of 82.15%. By adding demographics information as an input into Siamese network, the performance variance increased significantly.

**Table 2.** Comparison of different models, preprocessing techniques, input types, and loss functions.

| Architecture | Preprocessing | Loss | Input | Accuracy (%) | F1 score (%) |
| --- | --- | --- | --- | --- | --- |
| MLP | ViT | Cross-entropy | Radiographs | $54.08 \pm 3.88$ | $39.86 \pm 4.63$ |
| CNN | ViT | Cross-entropy | Radiographs | $55.14 \pm 4.13$ | $43.34 \pm 3.08$ |
| Siamese | ViT | Cross-entropy | Radiographs | $84.23 \pm 14.40$ | $79.88 \pm 19.47$ |
| Siamese | ViT | Contrastive | Radiographs | $86.17 \pm 19.67$ | $81.71 \pm 27.15$ |
| Siamese | ViT | Contrastive | Radiographs +<br>Demographics | $86.32 \pm 1.60$ | $82.15 \pm 2.06$ |

As shown in Figure 2a, most of the model’s confusion happened between 1 *< time ≤*5 (true label) and *time >* 5 (prediction) (n=17), between 1 *< time ≤*5 (true label) and *time >* 5 (prediction) (n=15), and between 1 *< time≤* 5 (true label) and *time >* 5 (prediction) (n=14). The actual time span for all misclassified samples is shown in Figure 2b, reflecting significant number of misclassified samples around *year* = 1 especially for predicted class 1 *<time ≤* 5. Moreover, the predictions of classes 1 *<time≤*5 and *time >* 5 are more concentrated around fewer time intervals, whereas distribution of predictions of class *time ≥*1 exhibits higher dispersion, which is reasonable taking into account the duration of time categories.

**Figure 2.**
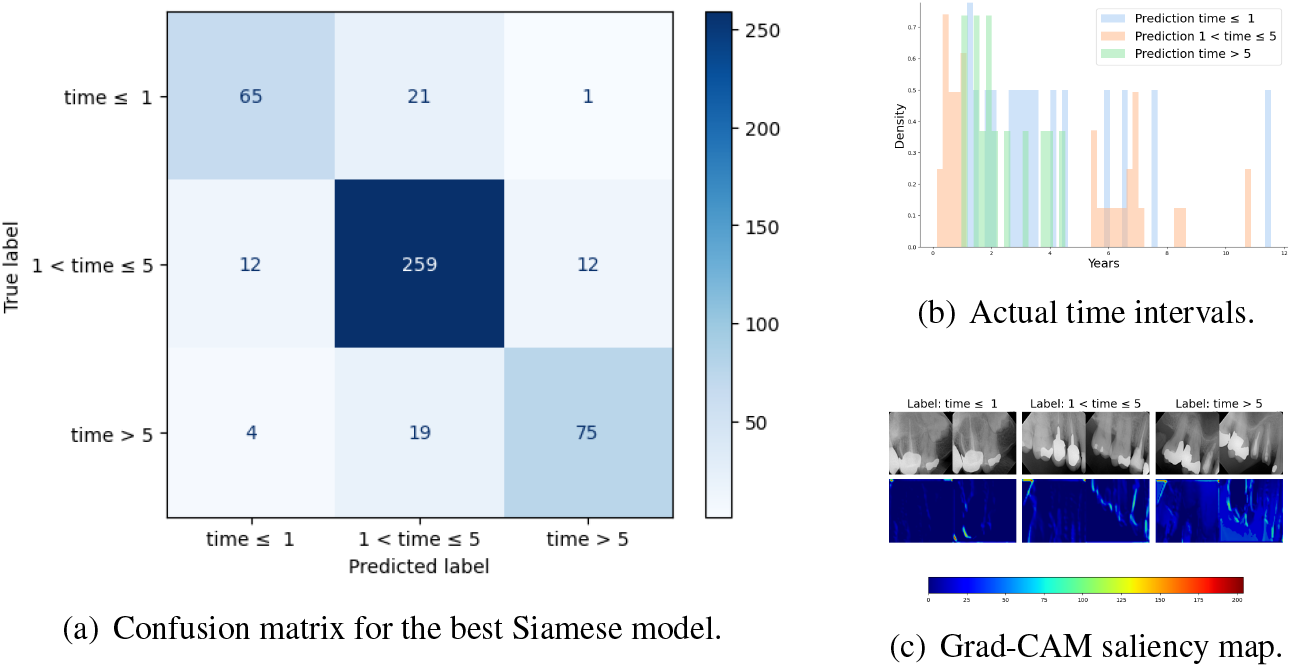
(a) Confusion matrix for the time interval categories and (b) Distribution of actual time intervals for all misclassified samples. (c) Visualization of parts of the input image where model’s dominantly focuses.

The focus of the Siamese model for pairs of input images for correctly classified samples is shown in Figure 2c. It highlights the importance of teeth shape as an indicator of time evolution and indicates that friction evolution and tooth wear could be used by the model to predict time span between pairs of radiographs [14]. However, the image corners also exhibit relatively high focus, which requires further investigation.

## 4. Conclusion

This study explored the use of Siamese networks to classify time intervals between dental visits based on periapical radiographs and patient demographic data. The novelty of this approach lies in the model’s ability to capture changes between radiographs, highlighting its potential to assist dental practitioners in estimating patient trajectories. In addition, we avoid the need for extensive and costly annotated datasets, while enabling large-scale generalization assessment of dental X-ray evolution models. Future work will focus on self-supervised learning models for dental disease prognosis, particularly in clinical settings where manually labeled data is unavailable.

## Data Availability

The data used in this study are not publicly available due to privacy and ethical restrictions.

